# Identifying gaps in health literacy research through parental participation

**DOI:** 10.1101/2023.06.15.23291427

**Authors:** Eva-Maria Grepmeier, Laura Pöhnl, Julia von Sommoggy, Maja Pawellek, Jonas Lander, Anja Alexandra Schulz, Claudia Hasenpusch, Anja Schwalfenberg, Marie-Luise Dierks, Eva Maria Bitzer, Christian Apfelbacher

## Abstract

**Introduction:** Involving patients and the public in design, conduct and dissemination of research has gained momentum in recent years. While methods to prioritize research on treatment uncertainties have been successfully applied for various disease entities, patient and public involvement has not been prominent to prioritize research in health literacy (HL). This study aimed to set up a participatory process on identifying HL research gaps from a parent’s perspective in two use cases: early childhood allergy prevention (ECAP) and COVID-19 in children with allergies (COVICAL).

**Methods:** To prepare and empower parents, we developed and provided preparatory webinars, introductory materials, i.e., factsheets and a brochure, and a scientific podcast with seven episodes. Recruitment was carried out by our cooperation partner German Allergy and Asthma Association e. V., via local day care centres and paediatricians as well as via snowballing. The identification of research gaps took place within five workshops with n= 55 participants, four face-to-face-workshops across Germany, one online workshop. Research ideas and needs were reviewed for overlap and redundancy and compared to the existing research state of the art.

**Results:** More than 150 initial research ideas and needs were collected which after review were reduced to a total of 37 ECAP, 33 COVICAL and 7 generic HL research questions. These were particularly related to the ease of finding and presenting good quality health information, information environment, health communication, professional education, and HL testing.

**Conclusions:** Involving parents in the formulation of HL research priorities proved to be challenging but feasible. Research ideas often reflect wishes directed at health professionals and the health system, i.e., organizational, and systemic HL. An e Delphi process will follow to elicit the TOP 10 research priorities in each use case. This project will help to plan patient/parent centred HL research in ECAP and COVICAL.

**Plain Language Summary:** We carried out a participatory HL research exercise to identify HL research gaps from a parent’s perspective. The aim was to learn about parents’/participants’ uncertainty in two use cases ECAP and COVICAL and what issues they would like to be answered by scientists. This resulted in a total of 37 ECAP, 33 COVICAL and 7 generic HL research questions.

## Introduction

Participatory approaches reflect and focus actual needs, knowledge and interests of patients, parents and citizens in general (Jilani et al., 2020). This may reduce “research waste” by focusing research on patient relevant issues through involvement (Buhr & Tannen, 2020; Chalmers & Glasziou, 2009). On an international level, health-related research with and by citizens is a common method to set research priorities, design, conduct, and disseminate research (Conklin, Morris, & Nolte, 2015; Domecq et al., 2014; Li, Abelson, Giacomini, & Contandriopoulos, 2015; Tritter & McCallum, 2006). In German-speaking countries participatory approaches became more important as well (International Collaboration for Participatory Health Research, o. J.; Ollenschläger, Wirth, Schwarz, Trifyllis, & Schaefer, 2018; Peter et al., 2020; Schilling et al., 2019; Wright, 2021). Involving patients in setting priorities for research has been successfully used regarding treatment uncertainties (Crowe, Fenton, Hall, Cowan, & Chalmers, 2015), in community settings (Breault et al., 2018; Clarke et al., 2023), and health promotion (Bush et al., 2017; Krawiec, Fisher, Du Toit, Bahnson, & Lack, 2021; Wright, 2021). Involvement and engagement with target groups increases the likelihood that health literacy (HL) interventions are effective (Batterham et al., 2014) but an approach to agree on research priorities has not yet been used in the field of HL research.

### Scope of the priority setting: Health Literacy in the use cases Early Childhood Allergy Prevention and COVID-19 in Children with Allergies

The German public health research group HELICAP “Health literacy in early childhood allergy prevention: parental competencies and public health in a shifting evidence landscape” explores health literacy (HL) in two use cases: early childhood allergy prevention (ECAP) and COVID-19 in children with allergies (COVICAL). HL is understood as the ability to access, understand, appraise, and apply health information to enable good health-related decisions (Sørensen et al., 2012). It is a multi-facetted concept determined by individual, situational and environmental factors (Abel & Sommerhalder, 2015; Bitzer & Sørensen, 2018; Sørensen et al., 2012). The goal of HL is to enable people to make grounded judgments, e.g., about when medical expert advice should be sought, knowledge of how to find the appropriate expert, and the ability to explain the health problem and personal concerns. In practice, it is much more difficult to act in a health-justified way as long as there is a lot of uncertainty about health issues. Uncertainty might induce anxiety and stress, further impeding good health-related decisions (Khojasteh, Davani, Shamsipour, Haghani, & Glamore, 2022; Okan et al., 2020; Schaeffer, Hurrelmann, Bauer, & Kolpatzik, 2018).

Both our use cases are characterized by high levels of uncertainty not least because of a changing evidence landscape: The scientific evidence on ECAP as well as recommendations by guidelines to prevent allergies in early childhood shifted from allergen avoidance (“protecting” the immune system) to early exposure (hence stimulation of the immune system) (Brough et al., 2022; Krawiec et al., 2021; Royal & Gray, 2020). In the first two years of the COVID-19 pandemic uncertainty was widespread among scholars, politicians, and the public. Rapidly emerging and changing scientific evidence hampered health literate decisions, a difficulty enhanced by the large amount of misinformation and unreliable information related to the pandemic (Borges do Nascimento et al., 2022; Okan et al., 2020; Schaefer, Bitzer, Okan, & Ollenschläger, 2021; Schaeffer et al., 2018).

The pandemic illustrated how important it is for individuals to be able especially to access, understand, appraise and apply health information (Abel & McQueen, 2020; Paakkari & Okan, 2020). For example in the case of people with allergic asthma, the risk assessment for COVID-19 infections was constrained by inconsistent scientific evidence: Asthma was initially considered to increase the risk for severe COVID-19 illness as respiratory viruses aggravate chronic airway diseases (Jackson et al., 2020). However, other studies suggested that asthma and respiratory allergies do not increase the risk for severe COVID-19 disease (Jackson et al., 2020; Wu & McGoogan, 2020).

Since both use cases enatil uncertainty further research into HL in relation to ECAP and COVICAL is warranted. Fostering HL concerning ECAP and COVICAL is an important public health concern, as e. g. low parental HL is linked to poorer health outcomes in (young) children, and lowers effectiveness in preventing disease in children (Buhr & Tannen, 2020; DeWalt, Dilling, Rosenthal, & Pignone, 2007; DeWalt & Hink, 2009; Miller, Lee, DeWalt, & Vann, 2010; Morrison, Glick, & Yin, 2019; Sanders, Shaw, Guez, Baur, & Rudd, 2009; Stafford, Goggins, Lathrop, & Haddad, 2021).

This study aimed to set up a participatory process to identify HL research gaps from a parent’s perspective in the fields of ECAP and COVICAL.

## Methods

The development and implementation of a participatory research process to identify research gaps from the parents’ perspective took place within the DFG-funded research group HELICAP - "Health literacy in early childhood allergy prevention: parental competencies and public health context in a shifting evidence landscape" (FOR 2959, project number: 409800133, spokesperson C.A. Apfelbacher).

Consisting of six work packages, HELICAP (www.helicap.org) adresses various research challenges:

(1) conflicts of interest in national and selected international ECAP-guidelines, (2) living systematic reviews on ECAP and COVID-19 related HL, (3) how health professionals translate available evidence into practice, (4) the degree to which health information on the internet meets parents’ needs, (5) factors influencing new parents’ HL and ECAP behaviours, (6) measurement of ECAP and COVID-19 related HL.

Representatives of each of the HELICAP work packages, of the HELICAP’s coordinating centre, and a patient representative from the German Allergy and Asthma Association (DAAB) formed a Task Force (TF), guiding the study process. The twelve TF-members have different scientific backgrounds (i.e., medicine, sociology, (health) educational sciences, cultural sciences, psychology, public health) and career levels (for details see appendix).

We started with a preparatory phase that included the development of introductory information, followed by interactive workshops to identify research gaps related to HL.

### Methodological Framework

Our study was guided by the James Lind Alliance’s Priority Setting Partnership framework (James Lind Alliance, 2021). This framework provides the fundamental basis of the planning, implementation, and evaluation of the participatory process. However, due to the scope and context of the study, we adapted the methodology to a framework for prioritization in the field of HL research. To support the integrity of our findings, we used both the REPRISE reporting framework (Tong et al., 2019) and the GRIPP2 checklist (Staniszewska et al., 2017) that have been developed to improve the reporting of patient and public involvement within a research study and associated publications.

### Target Group and Recruiting

The target group of this study were (new) parents with children, at average or high risk for atopic eczema, food allergies/anaphylaxis, or asthma/respiratory allergies, as well as with children already affected by allergies.

Recruitment took place during January-May and October-November 2022 via announcements on the DAAB and HELICAP website, through the DAAB newsletter and mailing lists, social media, as well as personal contacts and subsequent snowballing. At the HELICAP sites we approached parents in person and through employees or health professionals in family facilities, day-care centres, kindergartens, and paediatricians’ offices.

### Preparatory phase

To prepare and empower parents, we developed and provided webinars, introductory materials, i.e., factsheets and a brochure, and a scientific podcast.

- **Webinars**: We designed webinars based on each of the six HELICAP working packages, scheduled for 1.5 hours. The webinars followed a common guidance: 1) introducing briefly the HELICAP research fields and participatory research; 2) explaining the topic with regard to its meaning for and relevance towards parents; and 3) a discussion with the participants to give room for questions, ideas and comments. Each of the webinars was conducted twice to provide alternative time slots. In total we conducted 12 webinars during February-May 2022.
- **Factsheets**: Along with each webinar we provided brief digital factsheets. These factsheets with a mix of textual and visual information summarised the content of the webinar, contained room for participants’ ideas, notes, or questions, and offered references for further reading.
- **Brochure**: Based on the factsheets and the discussions at the webinars, we created a 16-page brochure as a single written plain language summary. The brochure includes questions and insights that emerged during the webinar discussion with participants and aimed at informing participants of the workshops to support their preparation.
- **Podcasts**: As both, the DAAB and parents who participated in the webinars repeatedly emphasized the importance of communicating the research project via a freely available audio format, we created a scientific podcast with seven episodes (again based on the six webinars, plus a general introductory episode). Each episode lasts about 20-35 minutes and is moderated by the DAAB representative.

All material is in German language and publicly available via the HELICAP website (HELICAP, 2022a, 2022b, 2022c), the podcasts also via an audio streaming service (Spotify, 2022).

### Interactive workshops to elicit research gaps

We conducted five interactive Workshops in October and November 2022 – four on-site in Regensburg, Hannover, Freiburg und Magdeburg and one online workshop – to identify which topics our target groups were missing that could help them make good health-related decisions for themselves and their children regarding ECAP and COVICAL.

To facilitate participation in the workshops, to provide a low-threshold access and create a feel-good atmosphere, we chose central locations used for family activities. The participants received a financial compensation, some workshops offered childcare, breakfast, or a bilingual implementation.

Workshops lasted about 2.5 hours (on-site) and 1.5 hours (digital), respectively. The time in the online workshop could be reduced because there was no need to physically move from table to table and the groups could spread out more quickly. In addition, the reduced time should also increase the willingness to participate.

#### Didactic concept

The workshop structure and methods encompassed participant orientation and constructivist didactics (Luchte 2012; Quilling 2015). These are particularly suitable for heterogeneous groups, allowing for aligning with participants’ needs, interests, and experiences (Quilling 2015). Depending on feedback from participants and any challenges in the process, minor adjustments were made from workshop to workshop, e.g. with regard to the material provided to participants. To identify the research gaps in HL in the two use cases rather than to communicate knowledge we had to stimulate participants to focus on how they deal with uncertainties and to reflect on what they do not know. Participants were given tasks and encouraged to review and critically question their own experiences and their factual knowledge about the use cases.

#### Structure

We organized the workshop as a moderated focus group discussion with four parts: introduction, main activity, summary, and conclusion (cf. table 1). The group-discussions were recorded, and the moderators took minutes in a supportive manner. The main activity covered three major topics:

a. “Accessing Health Information“: Reflecting own behaviour when searching online for a child health-related information.
b. “Measuring Health Literacy”: Identifying benefits and challenges when participating in a HL test from the participant perspective.
c. “Understanding Health Information“: Discussing the flow of information from health professionals to patients, understanding of how health information is evaluated, and how recommendations for action are applied to one’s life.

**Table 1:** Didactic structure of the HELICAP workshops to elicit research gaps with parents.

| Session | Goals | Description of session |  | Social format/<br>Methods | Material | Examples |
| --- | --- | --- | --- | --- | --- | --- |
| Introduction<br>(25 min) | Introduction of the study, the research area and getting to know each other | General information and formalities, information about the research group HELICAP, introduction of HL and the use cases, round of introductions |  | Plenary | PowerPoint slides, HELICAP brochure, consent form, privacy statement |  |
| Main activity<br>(85 min)<br>(3 x 25 min + 10 min break) | Reflecting on one's own search behaviour to derive limitations and challenges to access health information, and to elicit research needs. | Accessing Health Information | Activation: short internet search of COVID-19 in children with allergies. | Focus group discussion, think-aloud | Notebooks, tablet, smartphones, flash cards | Do you feel confident to search the Internet and deal with search results?<br>During your search, what questions did you have? |
|  |  |  | Discussion: Reflecting on challenges, difficulties, and limitations of searching for health information on the internet. |  |  |  |
|  |  |  | Deriving ideas: Collecting ideas for future research on the topic of accessing health information on the internet. |  |  |  |
|  | Find out when and how measurement of HL is useful and identify the added value of HL measurement (for individual, treatment, etc.) to elicit research needs. | Measuring Health Literacy | Activation: Reading and completing various health literacy tests. | Focus group discussion, think-aloud | Various health literacy tests, flash cards | What do you think of the questionnaires, what did you feel when you filled them in? In what situations might it be useful to complete such a questionnaire? |
|  |  |  | Discussion: Find out the circumstances for taking part in a HL test, what advantages/disadvantages HL tests offer from the participants point of view, and when parents need to know their HL. |  |  |  |
|  |  |  | Deriving ideas: Collecting ideas for future research on the topic of health literacy measurement |  |  |  |
|  | What are the problems in understanding health information, how are they dealt with, and how can research contribute to making information (transfer) more understandable? | Understanding Health Information | Activation: Reading a text on evidence shift in ECAP. | Focus group discussion, think-aloud | Sample texts on evidence shift, flash cards | How do you make sure you have understood something, and what do you do when you have a misunderstanding? What can be done to improve understanding of health information? |
|  |  |  | Discussion: Flow of information from health professionals or the media to patients, understanding of health information and how to deal with it. |  |  |  |
|  |  |  | Deriving ideas: Collecting ideas for future research on the topic of understanding health information |  |  |  |
| 15 min break |  |  |  |  |  |  |
| Summary<br>(20 min) | Placement of the results in the overall context, checking the completeness of the ideas | Presentation of the items collected at each major topic, possibility for additions and requests |  | Plenary | Items written down on flash cards |  |
| Conclusion<br>(10 min) | Thanks and Goodbye, building participant loyalty for further study steps | Last formalities, presentation of the next steps of the study |  | Plenary | Form for reimbursement of expenses |  |

During the main activity, we followed a procedure that allowed all participants to elaborate on each of the three topics. At the beginning, participants formed small groups with three to five persons. The small groups rotated through the three topics with 25 minutes time to deliberate on each, supported by a moderator. In the online workshop, we reduced the time to work on each topic to 15 minutes.

The units were designed as follows: First, participants were given a short task:

a. to search the internet on the topic of COVID-19 in children with allergies
b. to look at or try out different HL tests (e. g. HLSEU-Q16, Berlin Numeracy Test, CHC-Szenario 1, S-TOFHLA, and HELICAP questionnaire)
c. to read a text which addresses the evidence shift in allergy prevention.

Second, participants reflected on their experiences performing this task, started to discuss research needs, and made notes. Finally, the moderator summarized the collected ideas on cards and presented them on board to ensure that completeness.

#### Analysis and data synthesis

In the first workshop it turned out that formulating research gaps in term of precise questions is challenging for parents. We therefore extended the identification of research gaps to the collection of uncertainties, questions, and needs (in the following, referred to as “ideas for research”). After removing duplicates, we sorted the ideas derived in the five workshops, and translated the findings into potential research questions.

The discussions during the different sessions were not always strictly focused, despite the efforts of the moderators. There was some overlap in the topics and new topics were raised by the parents. After a review of the material, it was therefore necessary to expand the original three topics of the units (accessing health information, measuring HL, understanding health information) to a total of five categories, into which the ideas were inductively divided: "Health Information", "Information Environment", "Health Communication", “Professional Education", and "Health Literacy Testing".

To achieve a uniform level of abstraction and complexity the members of the TF reformulated the individual ideas in a two-stage process into scientific research questions and reviewed each individual aspect for overlap and redundancy.

In addition, each research question was assigned a unique identification number (ID). Wherever applicable, general phrased ideas were put into research questions with reference to ECAP and/or COVICAL. We focused on research questions for which subject-matter expertise exists in the HELICAP research group. Data processing was done using MAXQDA and Microsoft Excel.

Participants subsequently received an initial results overview via email, which is available at the HELICAP homepage (HELICAP, 2022d) and information about the further course of the study.

**Table 2:** Exemplary workflow of summarizing and collating research gaps.

| ID | Workshop theme | Parents ideas and needs | Category | Development of an initial question | Revising of question | Research question related to ECAP and COVICAL |
| --- | --- | --- | --- | --- | --- | --- |
| 5 | Accessing Health Information | Trust marks for websites, trustmarks for good websites, Certificate, Emblem for websites - "approved" by a reputable source. "Interdisciplinary group of experts", Help in assessing the seriousness of websites (also via radio, television, etc.) | Health Information | What can parents do to assess the reliability of health information on the internet, and how do they do it? | How can parents check the reliability and trustworthiness of health information on the internet? How can they do this? | How could parents check health information on the internet on ECAP for reliability and trustworthiness, and how do they do it actually? |
|  |  |  |  |  |  | How could parents check health information on the internet on COVICAL for reliability and trustworthiness, and how do they do it actually? |
| 19 | Understanding Health Information | Peer to Peer | Health Communication | What are the advantages of a target group-specific "peer to peer" mediation of health information? | What are the benefits of peer to peer exchange of health information? | What are the advantages of a targeted "peer to peer" exchange on ECAP? |
|  |  |  |  |  |  | What are the advantages of a targeted "peer to peer" exchange on COVICAL? |
| 39 | Measuring Health Literacy | Colours, graphs, pictures, tables simplify a test (visualise)<br>Time problem: tests should not take long<br>Pro "tick test": there is no/right and wrong<br>Questionnaire should not be too long (10-15 min.) | Health Literacy Testing | To what extent do the characteristics of a test, in particular the answer format, the preparation and the length of the test, influence willingness to participate and motivation in HL tests? How can this be put into practice? | To what extent do the characteristics of a test (e.g. answer format, preparation, test length) influence the willingness and motivation to take a particular HL test? | To what extent do the characteristics of a test (i.e. measurement approach, answering format, presentation and length of the test) influence the willingness and motivation to participate in HL tests on ECAP? |
|  |  |  |  |  |  | To what extent do the characteristics of a test (i.e. measurement approach, answering format, presentation and length of the test) influence the willingness and motivation to participate in HL tests on COVICAL? |

## Results

### Participants characteristics

A total of n=55 parents attended the workshops (on-site: n=45, online: n=10), 46 provided sociodemographic details. Participating parents were mostly female (78.3%), on average 38.7 years of age (SD=6.7). Educational attainment was high: 54,3% of the participants had a high school degree. 50% of the participants were affected by allergies, mostly pollen/gras. 41,3 % parents had an allergy affected child, with food allergy being most often mentioned (see Table 3).

**Table 3:** Characterization of workshop participants (n=46)

| Gender, n (%) |  | Years of education, n (%) |  |
| --- | --- | --- | --- |
| - female | 36 (78.3) | - high school | 34 (54.3) |
| - male | 10 (21.7) | - middle school | 10 (15.2) |
| Age in years (SD) | 38.7 (SD=6.7) | - others | 2 (10.9) |
| Suffering from allergy, n (%) | 23 (50) | Have a child with allergies, n (%) | 19 (41.3) |
| - grass and pollen | 11 (23.9) | - food allergies | 10 (21.7) |
| - animal hair | 5 (10.9) | - grass and pollen | 6 (13.0) |
| - house dust mite | 4 (8.7) | - house dust mite | 3 (6.5) |

### Parents view on research gaps

In total, we collected 152 ideas indicating research gaps in the workshops. These first were divided into the three broad workshop theme topic areas as follows: *Accessing Health Information* n= 61, *Measuring Health Literacy* n= 49, and *Understanding Health Information* n= 42.

After reviewing the material, the following five categories could be extracted:

<u>Health Information</u>: Participants expressed their desire that access to reliable health information (on the internet) needs to be much easier, for example by linking trustworthy websites or a barrier-free positioning of prevention topics. A comprehensible, target group-specific and multimedia presentation of health information was also described as desirable.

> ID27/28 “Support with research on the internet”, ID8 “Participatory development of brochures - citizens’ council for the production of health information”, ID5 “Seal, certificate, emblem for good websites - "approved" by reputable source - support in assessing the seriousness of websites (also via radio, television etc.), ranking the quality of information”

<u>Information Environment:</u> The parents clearly expressed the wish for a better exchange with other parents, e.g., on prevention issues. Paediatricians offer information about institutional contact points but cannot establish contact with other parents.

> ID10 “How can we encourage more exchange with other parents?”, ID11 “Which important information provider(s) should I know?”

<u>Health Communication:</u> Participants complained in general about the limited time of health professionals and information flow. Some participants indicated that from their experience physicians do not always try to provide medical information understandably to a layperson. Some would have liked to see health professionals use communication methods, such as "teach back". Others saw no need or too many barriers to implementing them. How to find out what you do not know so you can then search for information in a structured way was also mentioned.

> ID14 “Which professional group is most qualified to pass on information?”, ID23 “Framework: Empathy/Sensitivity/Space and Time/ Atmosphere” and ID21/22 “How can the teach-back method be established/improved?”

It became clear that there is a great need to share experiences with other parents or stakeholders. In addition, content on health topics should be communicated earlier, e.g., in daycare centers, kindergartens and schools.

> ID13 “How can we promote the exchange of experiences between those affected? (Promoting personal responsibility)", ID24 “How can parents be made aware of relevant topics?”

<u>Professional Education:</u> Participants wanted their children to be exposed to health literacy-related content at an early age. The question arose as to how this can be achieved in schools and kindergartens.

> ID26 “Integrate allergy prevention in the training of teachers and educators. Health literacy as a subject in school → Curriculum for school; Health literacy curriculum for kindergartens”

<u>Health Literacy Testing:</u> Research gaps in this topic highlight potential benefit and harms of HL testing. Parents wanted research to explore if testing for individual HL improves person-oriented and needs-based counselling. They wanted to know if self-testing for HL helps to better navigate the internet for health resources. Participants expressed concern about possible stigmatisation by a health professional if patients achieve a low HL level. They also reasoned about possible / optimal conditions to conduct HL-Tests.

> ID35: “Self-assessment: How good am I in the subject?”, ID38 “Test causes stigmatization and unequal treatment by doctors/health professionals” and ID46 “Preliminary consultations with a trained assistant better than an anonymous test situation!“

### Research Questions

The parental ideas, needs and questions resulted in a total of 45 research questions. Most of those research questions address both ECAP and COVICAL (n=32). Five research questions address ECAP only, one addresses COVICAL only. Seven questions do not explicitly refer to either ECAP or COVICAL, but are directed to more general issues, such as “How can HL be integrated best into the curriculum of schools?”. This means we collected a total of 37 research questions for ECAP, 33 for COVICAL and 7 generic HL questions.

Table 4 shows the finally derived research questions collected in the participatory process for the two use cases.

**Table 4:**
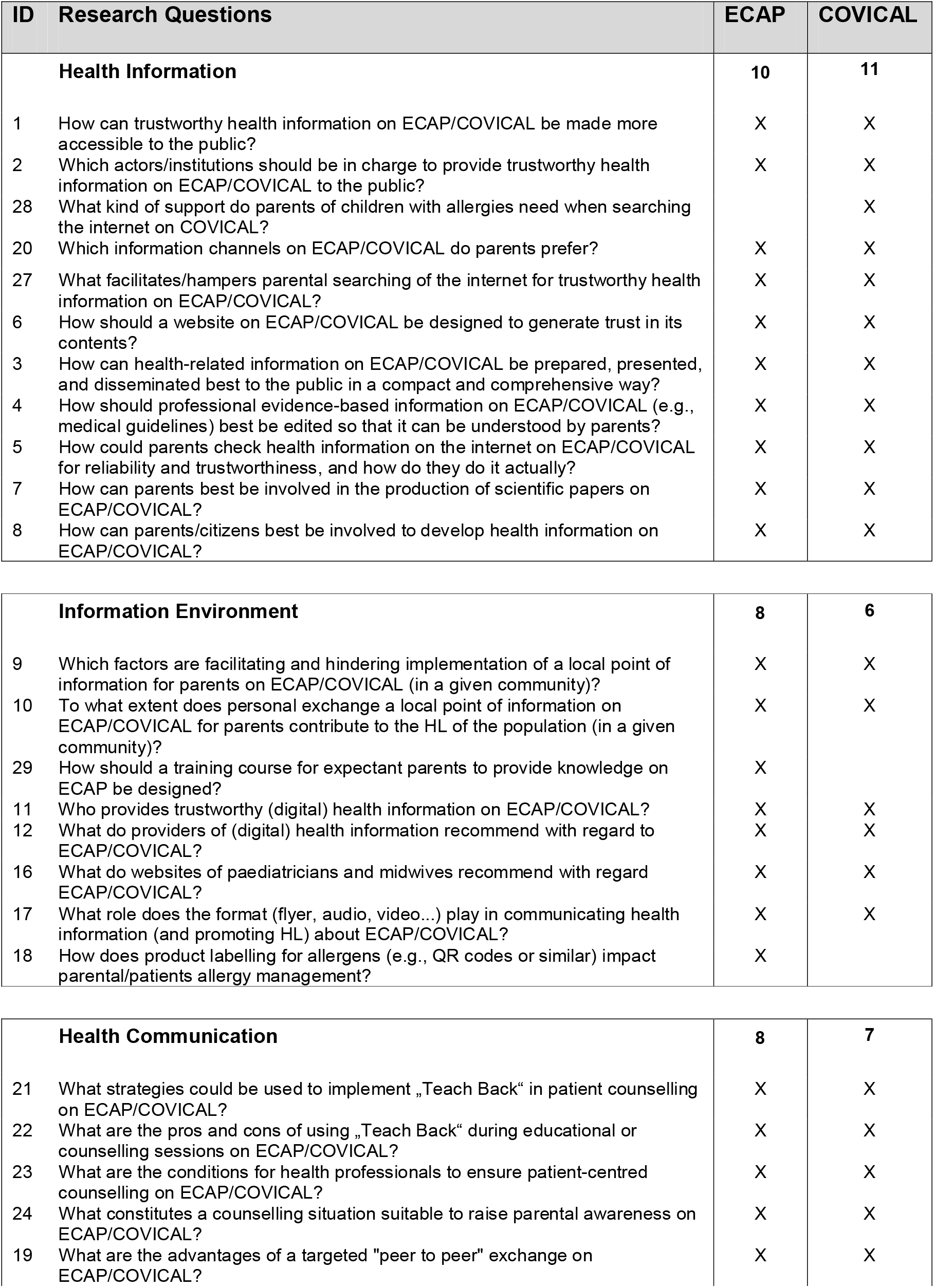
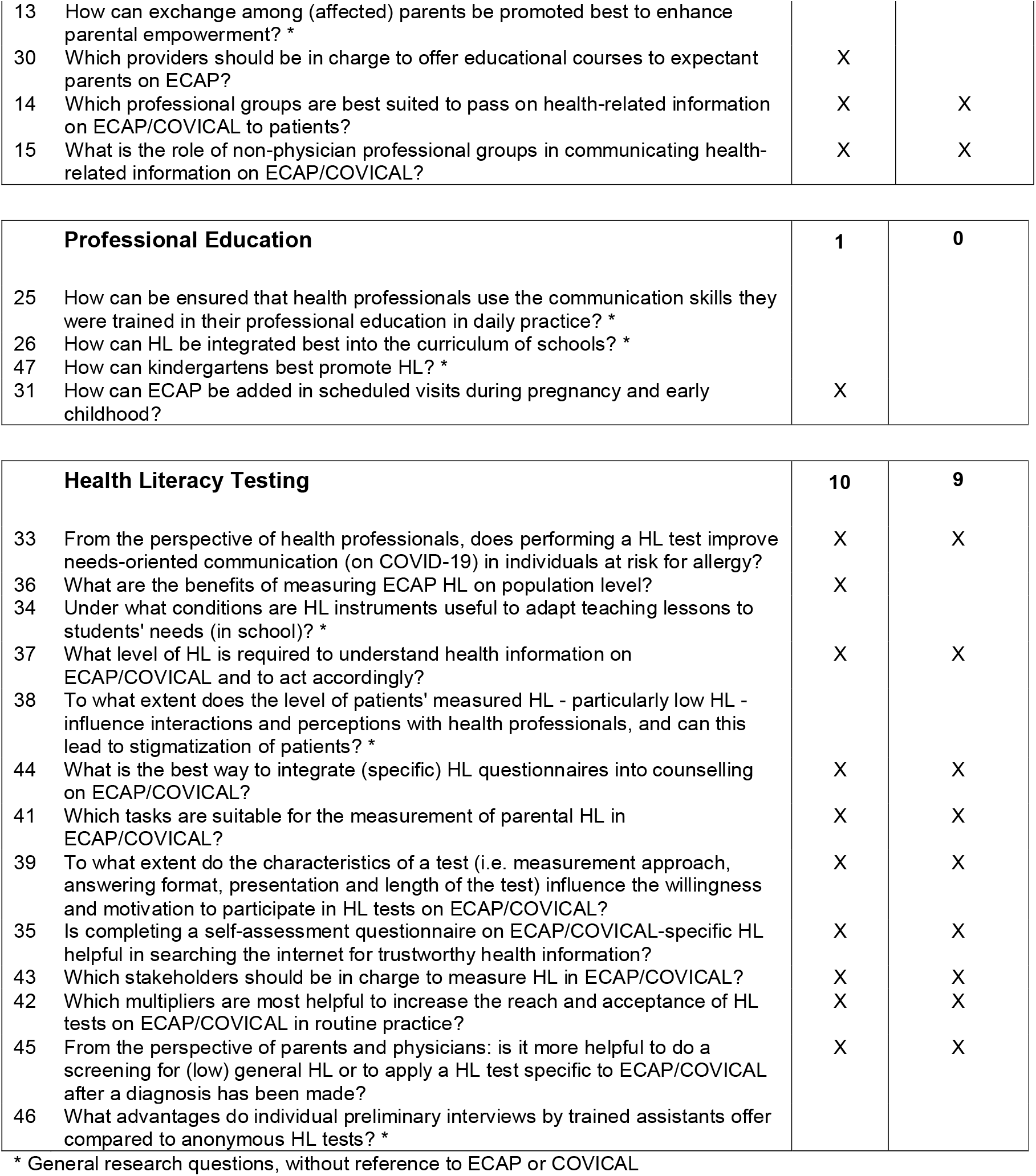
Research questions and allocation to use cases.

| ID | Research Questions | ECAP | COVICAL |
| --- | --- | --- | --- |
| **Health Information** |  | **10** | **11** |
| 1 | How can trustworthy health information on ECAP/COVICAL be made more accessible to the public? | X | X |
| 2 | Which actors/institutions should be in charge to provide trustworthy health information on ECAP/COVICAL to the public? | X | X |
| 28 | What kind of support do parents of children with allergies need when searching the internet on COVICAL? |  | X |
| 20 | Which information channels on ECAP/COVICAL do parents prefer? | X | X |
| 27 | What facilitates/hampers parental searching of the internet for trustworthy health information on ECAP/COVICAL? | X | X |
| 6 | How should a website on ECAP/COVICAL be designed to generate trust in its contents? | X | X |
| 3 | How can health-related information on ECAP/COVICAL be prepared, presented, and disseminated best to the public in a compact and comprehensive way? | X | X |
| 4 | How should professional evidence-based information on ECAP/COVICAL (e.g., medical guidelines) best be edited so that it can be understood by parents? | X | X |
| 5 | How could parents check health information on the internet on ECAP/COVICAL for reliability and trustworthiness, and how do they do it actually? | X | X |
| 7 | How can parents best be involved in the production of scientific papers on ECAP/COVICAL? | X | X |
| 8 | How can parents/citizens best be involved to develop health information on ECAP/COVICAL? | X | X |
| **Information Environment** |  | **8** | **6** |
| 9 | Which factors are facilitating and hindering implementation of a local point of information for parents on ECAP/COVICAL (in a given community)? | X | X |
| 10 | To what extent does personal exchange a local point of information on ECAP/COVICAL for parents contribute to the HL of the population (in a given community)? | X | X |
| 29 | How should a training course for expectant parents to provide knowledge on ECAP be designed? | X |  |
| 11 | Who provides trustworthy (digital) health information on ECAP/COVICAL? | X | X |
| 12 | What do providers of (digital) health information recommend with regard to ECAP/COVICAL? | X | X |
| 16 | What do websites of paediatricians and midwives recommend with regard ECAP/COVICAL? | X | X |
| 17 | What role does the format (flyer, audio, video...) play in communicating health information (and promoting HL) about ECAP/COVICAL? | X | X |
| 18 | How does product labelling for allergens (e.g., QR codes or similar) impact parental/patients allergy management? | X |  |
| **Health Communication** |  | **8** | **7** |
| 21 | What strategies could be used to implement „Teach Back“ in patient counselling on ECAP/COVICAL? | X | X |
| 22 | What are the pros and cons of using „Teach Back“ during educational or counselling sessions on ECAP/COVICAL? | X | X |
| 23 | What are the conditions for health professionals to ensure patient-centred counselling on ECAP/COVICAL? | X | X |
| 24 | What constitutes a counselling situation suitable to raise parental awareness on ECAP/COVICAL? | X | X |
| 19 | What are the advantages of a targeted "peer to peer" exchange on ECAP/COVICAL? | X | X |
| 13 | How can exchange among (affected) parents be promoted best to enhance parental empowerment? * |  |  |
| 30 | Which providers should be in charge to offer educational courses to expectant parents on ECAP? | X |  |
| 14 | Which professional groups are best suited to pass on health-related information on ECAP/COVICAL to patients? | X | X |
| 15 | What is the role of non-physician professional groups in communicating health-related information on ECAP/COVICAL? | X | X |
| **Professional Education** |  | **1** | **0** |
| 25 | How can be ensured that health professionals use the communication skills they were trained in their professional education in daily practice? * |  |  |
| 26 | How can HL be integrated best into the curriculum of schools? * |  |  |
| 47 | How can kindergartens best promote HL? * |  |  |
| 31 | How can ECAP be added in scheduled visits during pregnancy and early childhood? | X |  |
| **Health Literacy Testing** |  | **10** | **9** |
| 33 | From the perspective of health professionals, does performing a HL test improve needs-oriented communication (on COVID-19) in individuals at risk for allergy? | X | X |
| 36 | What are the benefits of measuring ECAP HL on population level? | X |  |
| 34 | Under what conditions are HL instruments useful to adapt teaching lessons to students' needs (in school)? * |  |  |
| 37 | What level of HL is required to understand health information on ECAP/COVICAL and to act accordingly? | X | X |
| 38 | To what extent does the level of patients' measured HL - particularly low HL - influence interactions and perceptions with health professionals, and can this lead to stigmatization of patients? * |  |  |
| 44 | What is the best way to integrate (specific) HL questionnaires into counselling on ECAP/COVICAL? | X | X |
| 41 | Which tasks are suitable for the measurement of parental HL in ECAP/COVICAL? | X | X |
| 39 | To what extent do the characteristics of a test (i.e. measurement approach, answering format, presentation and length of the test) influence the willingness and motivation to participate in HL tests on ECAP/COVICAL? | X | X |
| 35 | Is completing a self-assessment questionnaire on ECAP/COVICAL-specific HL helpful in searching the internet for trustworthy health information? | X | X |
| 43 | Which stakeholders should be in charge to measure HL in ECAP/COVICAL? | X | X |
| 42 | Which multipliers are most helpful to increase the reach and acceptance of HL tests on ECAP/COVICAL in routine practice? | X | X |
| 45 | From the perspective of parents and physicians: is it more helpful to do a screening for (low) general HL or to apply a HL test specific to ECAP/COVICAL after a diagnosis has been made? | X | X |
| 46 | What advantages do individual preliminary interviews by trained assistants offer compared to anonymous HL tests? * |  |  |

\* General research questions, without reference to ECAP or COVICAL

## Discussion

The present study used a participatory approach to identify gaps in HL research on health information, information environment, professional education and HL-testing in the fields of early childhood allergy prevention (ECAP) and COVID-19 on children with allergies (COVICAL).

We were able to show that the involvement of parents in the identification of HL research gaps is feasible.

Many of the ideas expressed by parents reveal a need for better health information and communication in routine preventive services and health care and thus address basic needs. However, research on accessibility, presentation, preparation, and quality of health information has been conducted for a long time. There are well validated checklists and criteria to identify credible health information on the internet (HoNCode; Institute for Quality and Efficiency in Health Care, 2021) and effective interventions to improve health communication are readily available (Centrella-Nigro & Alexander, 2017; Institute for Healthcare Improvement; Prochnow, Meiers, & Scheckel, 2019)(Michalopoulou, Falzarano, Arfken, & Rosenberg, 2010; UnityPoint Health, o. J.). Future research might not address those issues again but rather focus on how to find the most effective ways to implement such interventions at population level or in the health care system. It appears that there are not knowledge but rather implementation deficits in many areas pertaining to parental HL.

There are similar cases where research priorities set by citizens in fact reveal (basic) needs of the population, that are not resolved by further research but by political, governmental or health system targeted actions (Clarke et al., 2023; Trezona, Dodson, & Osborne, 2017).

Recruiting parents to the participatory process to identify HL research gaps on ECAP, and COVICAL proved to be challenging in our study.

One reason was that HL was not readily understood by our target group. Parents were not familiar with neither the expression nor the concept of HL and HL research. This became apparent during the workshops but also in the preparatory webinars, despite our efforts with a variety of different preparatory materials and a short thematic introduction at the beginning of the workshops; and despite a relatively high average level of education (54.3% of participants with high school diploma).

The specific topics – determined by the focus of our research group – might have further hampered the recruitment of parents: Often participatory approaches identifying research gaps are carried out with people directly affected by a condition, disease or health problem. Our first use case ECAP addresses primary prevention of allergies in children at average or high risk that do not suffer from allergies so far. In spite of the high and rising prevalence of allergies (Augustin et al., 2013; Böcking, Renz, & Pfefferle, 2012; Brough et al., 2022; Dierick et al., 2020). The topic appears not to be that salient in the population to be of major concern. Often parents become aware of childhood allergies only when the problem occurs. That is reflected in the composition of our sample, where 41.3% of the parents had children with manifest allergies, sometimes highly allergic. Those parents expressed some kind of “remorse” (“if I would have known that earlier”) reported on their very difficult journey to find relevant and trustworthy information and health care providers and were motivated to participate in the workshops because they wanted to “help other parents in future”. Under that assumption, for COVICAL it should have been easier to recruit parents. However, this was not the case, partly due to the fact, that at the time of recruitment (year 2022) the question on how to deal with COVID-19 in children with allergies/asthma was not that important any longer. A third reason might be that our participatory approach is not linked to a specific region, institution, or community and does not have solving the problem by developing an intervention in focus. In contrast to the Australian Optimizing Health Literacy and Access (OPHELIA) process (Ophelia, 2022), our study is not designed to put HL-interventions into practice. Where OPHELIA seeks answers to two key questions ‘What are the HL strengths and weaknesses of clients of participating sites?’ and ‘How do sites interpret and respond to these in order to achieve positive health and equity outcomes for their clients?’ and identifies, implements, and evaluates HL actions based on the needs of different stakeholders (Batterham et al., 2014; Ophelia, 2022), the participatory process described here aimed at eliciting research gaps in HL in two rather specific fields (ECAP, COVICAL).

Beyond the use cases, parents complained that information is often only provided when there is a perceived need from the perspective of health professionals - after the occurrence of symptoms. This is contrary to the idea of prevention, although research shows that counselling on preventive topics by health professionals can improve patients’, thus parents’ HL and health behaviour (Dennis et al., 2012; Gagliardi, Abdallah, Faulkner, Ciliska, & Hicks, 2015).

This study is based on an established method, which has been adapted and extended to our needs: the James Lind Alliance (JLA) approach. Contrary to the recommendation of the JLA (James Lind Alliance, 2021), we did not in advance search for evidence of uncertainty in documented sources of information and include this evidence in the process. A pre-testing for research gaps might have led to different results in our study and to participants refraining from formulating their basic needs, which is one of the key findings of the study. A systematic review of the outcomes and experiences of patient co-researchers shows that focus groups and individual group interviews are the most commonly used study designs. (Malterud & Elvbakken, 2020). We chose a focus group approach, which has been shown to have several advantages over interviews: New ideas that remain hidden or unrecognised in individual interviews can be stimulated by spontaneous utterances in the group, and, in larger groups, the gap between scientists and non-scientists might be smoothed out, fostering participation (Schulz, Mack, & Renn, 2012). Other studies used family workshops (Grabowski et al., 2022), observation and discussion (Scheffelaar et al., 2020) or online meetings with small group sessions (Ziegler et al., 2022) to identify stakeholder needs.

The experience gained from this study can help future similar initiatives. If we do not engage in co-production of research questions, we may fail to address the issues that matter most to the groups we want to benefit. Furthermore, participatory approaches could be beneficial in implementing research findings in the everyday life of beneficiaries. Involving parents could increase their understanding of HL research in ECAP and COVICAL. The research questions collected through the participatory process up to this point provide the basis for a subsequent Delphi process to identify the TOP 10 research questions in each of the two use cases. The overall aim of this process is to enable future research efforts in the field of HL on ECAP and COVICAL research to be focused and parent / patient-centred.

## Supporting information

GRIPP2 checklist (short form)

REPRISE framework

## Data Availability

All data produced in the present study are, if not contained in the manuscript, available upon reasonable request to the authors.

## Acknowledgement

The HELICAP Research Group would like to thank its cooperation partner, the German Allergy and Asthma Association (DAAB e.V.), and all participants in the study for their valuable contribution.

## Abbreviations

DAAB: German Allergy and Asthma Association
HL: Health Literacy
TF: Task Force
JLA: James Lind Alliance
ECAP: Early Childhood Allergy Prevention
COVICAL: COVID-19 In Children with Allergies

## Declarations

### Ethics approval and consent to participate

Participation in the entire study was voluntary and could be discontinued at any time without giving reasons. The study was reviewed and positively approved by the Ethics Committee of the Otto-von Guericke-University Magdeburg.

### Availability of data and materials

All data generated or analysed during this study are included in this published article and its supplementary information files

## Appendix

**Table 5:**
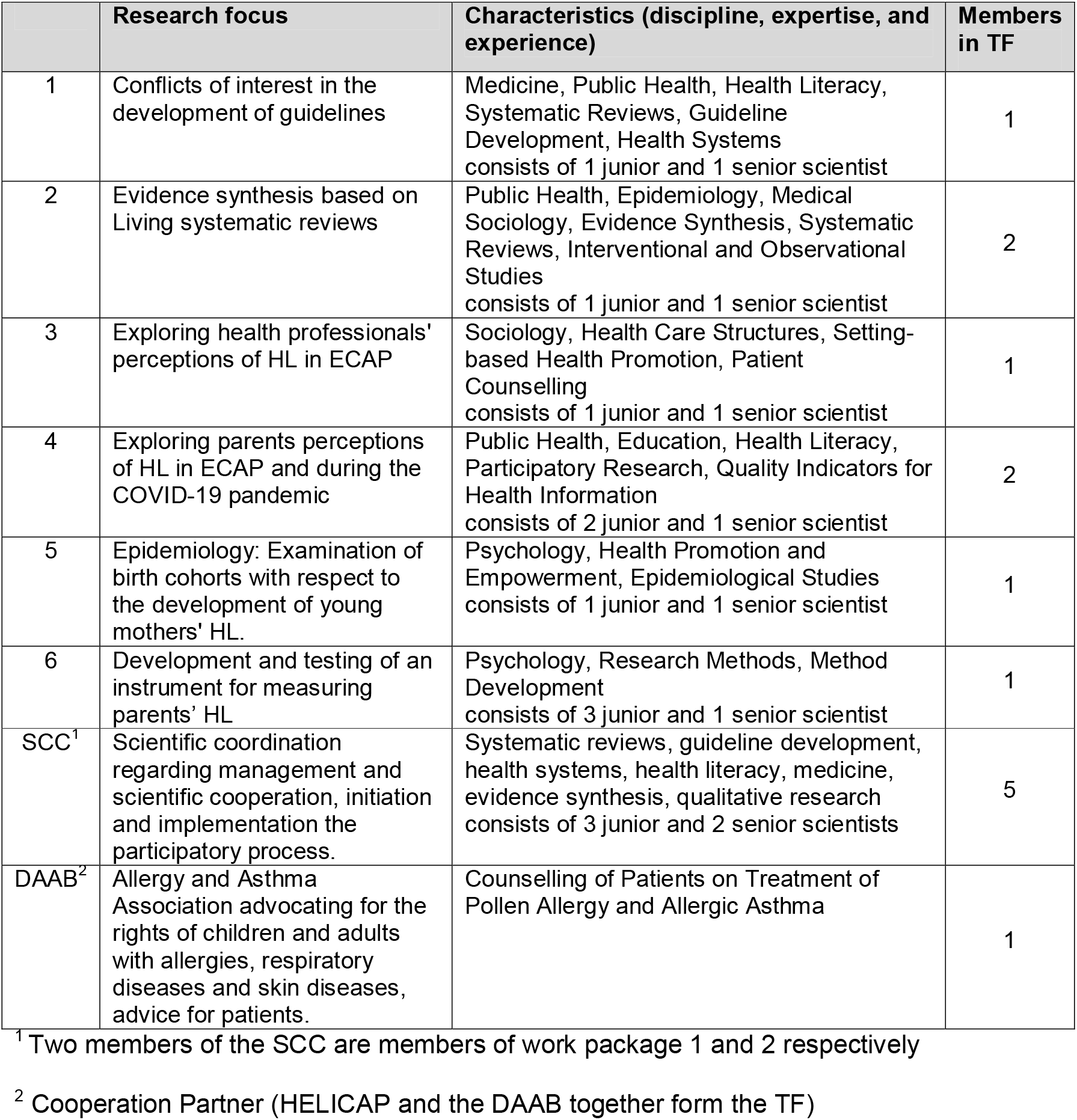
Characteristic of the Task Force (TF)

|  | Research focus | Characteristics (discipline, expertise, and experience) | Members in TF |
| --- | --- | --- | --- |
| 1 | Conflicts of interest in the development of guidelines | Medicine, Public Health, Health Literacy, Systematic Reviews, Guideline Development, Health Systems<br>consists of 1 junior and 1 senior scientist | 1 |
| 2 | Evidence synthesis based on Living systematic reviews | Public Health, Epidemiology, Medical Sociology, Evidence Synthesis, Systematic Reviews, Interventional and Observational Studies<br>consists of 1 junior and 1 senior scientist | 2 |
| 3 | Exploring health professionals' perceptions of HL in ECAP | Sociology, Health Care Structures, Setting-based Health Promotion, Patient Counselling<br>consists of 1 junior and 1 senior scientist | 1 |
| 4 | Exploring parents perceptions of HL in ECAP and during the COVID-19 pandemic | Public Health, Education, Health Literacy, Participatory Research, Quality Indicators for Health Information<br>consists of 2 junior and 1 senior scientist | 2 |
| 5 | Epidemiology: Examination of birth cohorts with respect to the development of young mothers' HL. | Psychology, Health Promotion and Empowerment, Epidemiological Studies<br>consists of 1 junior and 1 senior scientist | 1 |
| 6 | Development and testing of an instrument for measuring parents' HL | Psychology, Research Methods, Method Development<br>consists of 3 junior and 1 senior scientist | 1 |
| SCC<sup>1</sup> | Scientific coordination regarding management and scientific cooperation, initiation and implementation the participatory process. | Systematic reviews, guideline development, health systems, health literacy, medicine, evidence synthesis, qualitative research<br>consists of 3 junior and 2 senior scientists | 5 |
| DAAB<sup>2</sup> | Allergy and Asthma Association advocating for the rights of children and adults with allergies, respiratory diseases and skin diseases, advice for patients. | Counselling of Patients on Treatment of Pollen Allergy and Allergic Asthma | 1 |

<sup>1</sup> Two members of the SCC are members of work package 1 and 2 respectively
<sup>2</sup> Cooperation Partner (HELICAP and the DAAB together form the TF)

## References

Abel, T., & McQueen, D. (2020). Critical health literacy and the COVID-19 crisis. Health Promotion International, 35(6), 1612–1613. https://doi.org/10.1093/heapro/daaa040.

Abel, T., & Sommerhalder, K. (2015). Gesundheitskompetenz/Health Literacy: Das Konzept und seine Operationalisierung [Health literacy: An introduction to the concept and its measurement]. Bundesgesundheitsblatt - Gesundheitsforschung - Gesundheitsschutz, 58(9), 923–929. https://doi.org/10.1007/s00103-015-2198-2

Augustin, M., Franzke, N., Beikert, F. C., Stadler, R., Reusch, M., Schmitt, J., & Schäfer, I. (2013). Allergies in Germany - prevalence and perception by the public. Journal of the German Society of Dermatology : JDDG, 11(6), 514–520. https://doi.org/10.1111/j.1610-0387.2012.08049.x

Batterham, R. W., Buchbinder, R., Beauchamp, A., Dodson, S., Elsworth, G. R., & Osborne, R. H. (2014). The OPtimising HEalth LIterAcy (Ophelia) process: Study protocol for using health literacy profiling and community engagement to create and implement health reform. BMC Public Health, 14, 694. https://doi.org/10.1186/1471-2458-14-694

Bitzer, E. M., & Sørensen, K. (2018). Gesundheitskompetenz – Health Literacy [Health Literacy]. Das Gesundheitswesen, 80(08-09), 754–766. https://doi.org/10.1055/a-0664-0395

Böcking, C., Renz, H., & Pfefferle, P. I. (2012). Prävalenz und sozioökonomische Bedeutung von Allergien in Deutschland [Prevalence and socio-economic relevance of allergies in Germany]. Bundesgesundheitsblatt - Gesundheitsforschung - Gesundheitsschutz, 55(3), 303–307. https://doi.org/10.1007/s00103-011-1427-6

Borges do Nascimento, I. J., Pizarro, A. B., Almeida, J. M., Azzopardi-Muscat, N., Gonçalves, M. A., Björklund, M., & Novillo-Ortiz, D. (2022). Infodemics and health misinformation: A systematic review of reviews. Bulletin of the World Health Organization, 100(9), 544–561. https://doi.org/10.2471/BLT.21.287654

Breault, L. J., Rittenbach, K., Hartle, K., Babins-Wagner, R., Beaudrap, C. de, Jasaui, Y., . . . Mason-Lai, P. (2018). People with lived experience (PWLE) of depression: Describing and reflecting on an explicit patient engagement process within depression research priority setting in Alberta, Canada. Research Involvement and Engagement, 4, 37. https://doi.org/10.1186/s40900-018-0115-1

Brough, H. A., Lanser, B. J., Sindher, S. B., Teng, J. M. C., Leung, D. Y. M., Venter, C., . . . Nagler, C. R. (2022). Early intervention and prevention of allergic diseases. Allergy, 77(2), 416–441. https://doi.org/10.1111/all.15006

Buhr, E. de, & Tannen, A. (2020). Parental health literacy and health knowledge, behaviours and outcomes in children: A cross-sectional survey. BMC Public Health, 20(1), 1096. https://doi.org/10.1186/s12889-020-08881-5

Bush, P. L., Pluye, P., Loignon, C., Granikov, V., Wright, M. T., Pelletier, J.⍰F., . . . Repchinsky, C. (2017). Organizational participatory research: A systematic mixed studies review exposing its extra benefits and the key factors associated with them. Implementation Science: IS, 12(1), 119. https://doi.org/10.1186/s13012-017-0648-y

Centrella-Nigro, A. M., & Alexander, C. (2017). Using the Teach-Back Method in Patient Education to Improve Patient Satisfaction. Journal of Continuing Education in Nursing, 48(1), 47–52. https://doi.org/10.3928/00220124-20170110-10

Chalmers, I., & Glasziou, P. (2009). Avoidable waste in the production and reporting of research evidence. Lancet, 374(9683), 86–89. https://doi.org/10.1016/S0140-6736(09)60329-9

Clarke, E., Anderson-Saria, G., Kisoli, A., Urasa, S., Moloney, S., Safic, S., . . . Paddick, S.⍰M. (2023). Patient priority setting in HIV ageing research: Exploring the feasibility of community engagement and involvement in Tanzania. Research Involvement and Engagement, 9(1), 3. https://doi.org/10.1186/s40900-022-00409-y

Conklin, A., Morris, Z., & Nolte, E. (2015). What is the evidence base for public involvement in health-care policy? Results of a systematic scoping review. Health Expectations: An International Journal of Public Participation in Health Care and Health Policy, 18(2), 153–165. https://doi.org/10.1111/hex.12038

Crowe, S., Fenton, M., Hall, M., Cowan, K., & Chalmers, I. (2015). Patients’, clinicians’ and the research communities’ priorities for treatment research: There is an important mismatch. Research Involvement and Engagement, 1, 2. https://doi.org/10.1186/s40900-015-0003-x

Dennis, S., Williams, A., Taggart, J., Newall, A., Denney-Wilson, E., Zwar, N., . . . Harris, M. F. (2012). Which providers can bridge the health literacy gap in lifestyle risk factor modification education: A systematic review and narrative synthesis. BMC Family Practice, 13, 44. https://doi.org/10.1186/1471-2296-13-44

DeWalt, D. A., Dilling, M. H., Rosenthal, M. S., & Pignone, M. P. (2007). Low parental literacy is associated with worse asthma care measures in children. Ambulatory Pediatrics, 7(1), 25–31. https://doi.org/10.1016/j.ambp.2006.10.001

DeWalt, D. A., & Hink, A. (2009). Health literacy and child health outcomes: A systematic review of the literature. Pediatrics, 124(3), 265–274. https://doi.org/10.1542/peds.2009-1162B

Dierick, B. J. H., van der Molen, T., Flokstra-de Blok, B. M. J., Muraro, A., Postma, M. J., Kocks, J. W. H., & van Boven, J. F. M. (2020). Burden and socioeconomics of asthma, allergic rhinitis, atopic dermatitis and food allergy. Expert Review of Pharmacoeconomics & Outcomes Research, 20(5), 437–453. https://doi.org/10.1080/14737167.2020.1819793.

Domecq, J. P., Prutsky, G., Elraiyah, T., Wang, Z., Nabhan, M., Shippee, N., . . . Murad, M. H. (2014). Patient engagement in research: A systematic review. BMC Health Services Research, 14, 89. https://doi.org/10.1186/1472-6963-14-89

Gagliardi, A. R., Abdallah, F., Faulkner, G., Ciliska, D., & Hicks, A. (2015). Factors contributing to the effectiveness of physical activity counselling in primary care: A realist systematic review. Patient Education and Counseling, 98(4), 412–419. https://doi.org/10.1016/j.pec.2014.11.020

Grabowski, D., Pals, R. A. S., Hoeeg, D., Ingersgaard, M. V., DeCosta, P., & Jespersen, L. N. (2022). Participatory family workshops in psychosocial health and illness research: Experiences from Danish health promotion projects. Health Promotion International, 37(S2), ii73-ii82. https://doi.org/10.1093/heapro/daac014

HELICAP (2022a). Broschüre. Retrieved from https://www.helicap.org/eltern-machen-forschung/broschuere

HELICAP (2022b). Download Factsheets. Retrieved from https://www.helicap.org/webinar

HELICAP (2022c). Gesund und Kompetent: Der Podcast von und mit der Forschungsgruppe HELICAP in Kooperation mit dem Deutschen Allergie- und Asthmabund. Retrieved from https://www.helicap.org/eltern-machen-forschung/podcast

HELICAP (2022d). Workshop Ergebnisse: Zusammenfassung der Workshopergebnisse für die Eltern. Retrieved from https://www.helicap.org/fileadmin/helicap_content/PDF/Eltern_machen_Forschung_-_Workshopergebnisse.pdf)

HoNCode. Health On the Net: a non for profit organisation, promotes transparent and reliable health information online. Retrieved from https://www.hon.ch/en/

Institute for Healthcare Improvement. Ask Me 3: Good Questions for Your Good Health. Retrieved from https://www.ihi.org/resources/Pages/Tools/Ask-Me-3-Good-Questions-for-Your-Good-Health.aspx

Institute for Quality and Efficiency in Health Care (2021). Finding high-quality health information on the internet. Retrieved from https://www.informedhealth.org/finding-high-quality-health-information-on-the-internet.html

International Collaboration for Participatory Health Research (o. J.). What is Participatory Health Research (PHR)? Retrieved from http://www.icphr.org/

Jackson, D. J., Busse, W. W., Bacharier, L. B., Kattan, M., O’Connor, G. T., Wood, R. A., . . . 9Altman, M. C. (2020). Association of respiratory allergy, asthma, and expression of the SARS-CoV-2 receptor ACE2. The Journal of Allergy and Clinical Immunology, 146(1), 203-206.e3. https://doi.org/10.1016/j.jaci.2020.04.009.

James Lind Alliance (2021). The James Lind Alliance Guidebook: Version 10. Retrieved from https://www.jla.nihr.ac.uk/jla-guidebook/downloads/JLA-Guidebook-Version-10-March-2021.pdf

Jilani, H., Rathjen, K. I., Schilling, I., Herbon, C., Scharpenberg, M., Brannath, W., & Gerhardus, A. (2020). Handreichung zur Patient*innenbeteiligung an klinischer Forschung. Universität Bremen. https://doi.org/10.26092/ELIB/229

Khojasteh, D., Davani, E., Shamsipour, A., Haghani, M., & Glamore, W. (2022). Climate change and COVID-19: Interdisciplinary perspectives from two global crises. The Science of the Total Environment, 844, 157142. https://doi.org/10.1016/j.scitotenv.2022.157142

Krawiec, M., Fisher, H. R., Du Toit, G., Bahnson, H. T., & Lack, G. (2021). Overview of oral tolerance induction for prevention of food allergy-Where are we now? Allergy, 76(9), 2684–2698. https://doi.org/10.1111/all.14758

Li, K. K., Abelson, J., Giacomini, M., & Contandriopoulos, D. (2015). Conceptualizing the use of public involvement in health policy decision-making. Social Science & Medicine, 138, 14–21. https://doi.org/10.1016/j.socscimed.2015.05.023

Malterud, K., & Elvbakken, K. T. (2020). Patients participating as co-researchers in health research: A systematic review of outcomes and experiences. Scandinavian Journal of Public Health, 48(6), 617–628. https://doi.org/10.1177/1403494819863514

Michalopoulou, G., Falzarano, P., Arfken, C., & Rosenberg, D. (2010). Implementing Ask Me 3 to improve African American patient satisfaction and perceptions of physician cultural competency. Journal of Cultural Diversity, 17(2), 62–67.

Miller, E., Lee, J. Y., DeWalt, D. A., & Vann, W. F. (2010). Impact of caregiver literacy on children’s oral health outcomes. Pediatrics, 126(1), 107–114. https://doi.org/10.1542/peds.2009-2887

Morrison, A. K., Glick, A., & Yin, H. S. (2019). Health Literacy: Implications for Child Health. Pediatrics in Review, 40(6), 263–277. https://doi.org/10.1542/pir.2018-0027

Okan, O., Sombre, S. de, Hurrelmann, K., Berens, E.⍰M., Bauer, U., & Schaeffer, D. (2020). Gesundheitskompetenz der Bevölkerung im Umgang mit der Coronavirus-Pandemie. Bielefeld. Retrieved from Universität Bielefeld website: https://blogs.uni-bielefeld.de/blog/pressemitteilungen/resource/Gesundheitskompetenz_in_der_Coronavirus-Pandemie_2020_Endbericht.pdf

Ollenschläger, G., Wirth, T., Schwarz, S., Trifyllis, J., & Schaefer, C. (2018). Unzureichende Patientenbeteiligung an der Leitlinienentwicklung in Deutschland – eine Analyse der von der AWMF verbreiteten ärztlichen Empfehlungen [Patient involvement in clinical practice guidelines is poor after 12 years of German guideline standards: A review of guideline methodologies]. Zeitschrift für Evidenz, Fortbildung und Qualität im Gesundheitswesen, 135-136, 50–55. https://doi.org/10.1016/j.zefq.2018.06.006

Ophelia (2022). Ophelia (Optimising Health Literacy and Access). Retrieved from https://healthliteracydevelopment.com/

Paakkari, L., & Okan, O. (2020). COVID-19: health literacy is an underestimated problem. The Lancet Public Health, 5(5), e249–e250. https://doi.org/10.1016/S2468-2667(20)30086-4

Peter, S. von, Bär, G., Behrisch, B., Bethmann, A., Hartung, S., Kasberg, A., . . . Wright, M. (2020). Partizipative Gesundheitsforschung in Deutschland – quo vadis? [Participatory research in Germany - quo vadis?]. Gesundheitswesen, 82(4), 328–332. https://doi.org/10.1055/a-1076-8078

Prochnow, J. A., Meiers, S. J., & Scheckel, M. M. (2019). Improving Patient and Caregiver New Medication Education Using an Innovative Teach-back Toolkit. Journal of Nursing Care Quality, 34(2), 101–106. https://doi.org/10.1097/NCQ.0000000000000342

Royal, C., & Gray, C. (2020). Allergy Prevention: An Overview of Current Evidence. Yale Journal of Biology and Medicine, 93(5), 689–698.

Sanders, L. M., Shaw, J. S., Guez, G., Baur, C., & Rudd, R. (2009). Health literacy and child health promotion: Implications for research, clinical care, and public policy. Pediatrics, 124 Suppl 3, S306–14. https://doi.org/10.1542/peds.2009-1162G

Schaefer, C., Bitzer, E. M., Okan, O., & Ollenschläger, G. (2021). Umgang mit Fehl- und Desinformationen in Medien: Eine Übersicht über aktuelle wissenschaftliche Erkenntnisse und Handlungsempfehlungen zum Umgang mit Fehl- und Desinformationen bei COVID-19. Hintergrundpapier. Retrieved from https://www.public-health-covid19.de/images/2021/Ergebnisse/20210902_Hintergrund_Fehlinformation_update.pdf

Schaeffer, D., Hurrelmann, K., Bauer, U., & Kolpatzik, K. (2018). National Action Plan Health Literacy. Promoting Health Literacy in Germany. Berlin: KomPart.

Scheffelaar, A., Bos, N., Jong, M. de, Triemstra, M., van Dulmen, S., & Luijkx, K. (2020). Lessons learned from participatory research to enhance client participation in long-term care research: A multiple case study. Research Involvement and Engagement, 6, 27. https://doi.org/10.1186/s40900-020-00187-5

Schilling, I., Behrens, H., Hugenschmidt, C., Liedtke, J., Schmiemann, G., & Gerhardus, A. (2019). Patient involvement in clinical trials: Motivation and expectations differ between patients and researchers involved in a trial on urinary tract infections. Research Involvement and Engagement, 5, 15. https://doi.org/10.1186/s40900-019-0145-3.

Schulz, M., Mack, B., & Renn, O. (Eds.) (2012). Fokusgruppen in der empirischen Sozialwissenschaft: Von der Konzeption bis zur Auswertung. Wiesbaden: Springer VS. Retrieved from http://ebooks.ciando.com/book/index.cfm/bok_id/328734

Sørensen, K., van den Broucke, S., Fullam, J., Doyle, G., Pelikan, J., Slonska, Z., & Brand, H. (2012). Health literacy and public health: A systematic review and integration of definitions and models. BMC Public Health, 12, 80. https://doi.org/10.1186/1471-2458-12-80.

Spotify (2022). Gesund und Kompetent: HELICAP. Retrieved from https://open.spotify.com/show/0eA8GeGCyPFLqV94P8eNCA

Stafford, J. D., Goggins, E. R., Lathrop, E., & Haddad, L. B. (2021). Health Literacy and Associated Outcomes in the Postpartum Period at Grady Memorial Hospital. Maternal and Child Health Journal, 25(4), 599–605. https://doi.org/10.1007/s10995-020-03030-1

Staniszewska, S., Brett, J., Simera, I., Seers, K., Mockford, C., Goodlad, S., . . . Tysall, C. (2017). Gripp2 reporting checklists: Tools to improve reporting of patient and public involvement in research. BMJ (Clinical Research Ed.), 358, j3453. https://doi.org/10.1136/bmj.j3453

Tong, A., Synnot, A., Crowe, S., Hill, S., Matus, A., Scholes-Robertson, N., . . . Craig, J. C. (2019). Reporting guideline for priority setting of health research (REPRISE). BMC Medical Research Methodology, 19(1), 243. https://doi.org/10.1186/s12874-019-0889-3

Trezona, A., Dodson, S., & Osborne, R. H. (2017). Development of the organisational health literacy responsiveness (Org-HLR) framework in collaboration with health and social 8services professionals. BMC Health Services Research, 17. 8 https://doi.org/10.1186/s12913-017-2465-z

Tritter, J. Q., & McCallum, A. (2006). The snakes and ladders of user involvement: Moving beyond Arnstein. Health Policy (Amsterdam, Netherlands), 76(2), 156–168. https://doi.org/10.1016/j.healthpol.2005.05.008.

UnityPoint Health (o. J.). Always Use Teach Back! Retrieved from https://www.ihi.org/resources/Pages/Tools/AlwaysUseTeachBack!aspx

Wright, M. T. (2021). Partizipative Gesundheitsforschung: Ursprünge und heutiger Stand [Participatory health research: origins and current trends]. Bundesgesundheitsblatt - Gesundheitsforschung - Gesundheitsschutz, 64(2), 140–145. https://doi.org/10.1007/s00103-020-03264-y

Wu, Z., & McGoogan, J. M. (2020). Characteristics of and Important Lessons From the Coronavirus Disease 2019 (COVID-19) Outbreak in China: Summary of a Report of 72 314 Cases From the Chinese Center for Disease Control and Prevention. JAMA, 323(13), 1239–1242. https://doi.org/10.1001/jama.2020.2648

Ziegler, S., Raineri, A., Nittas, V., Rangelov, N., Vollrath, F., Britt, C., & Puhan, M. A. (2022). Long COVID Citizen Scientists: Developing a Needs-Based Research Agenda by Persons Affected by Long COVID. The Patient, 15(5), 565–576. https://doi.org/10.1007/s40271-022-00579-7

