## Supplementary material for "Identifying gaps in health literacy research through parental participation": GRIPP2 checklist (short form)

GRIPP 2 Checklist / Short form

| **Section and topic** | **Item** | **Reported on page No**  **(Page numbers refer to the submitted Word document** |
| --- | --- | --- |
| 1: Aim | Report the aim of PPI in the study | Abstract: 3  Introduction: 7 |
| 2: Methods | Provide a clear description of the methods used for PPI in the study | Abstract: 3  Methods: 10-17 |
| 3: Study results | Outcomes—Report the results of PPI in the study, including both positive and negative outcomes | Abstract: 3  Results: 18-23 |
| 4: Discussion and conclusions | Outcomes—Comment on the extent to which PPI influenced the study overall. Describe positive and negative effects | Abstract: 3  Discussion: 24-27 |
| 5: Reflections/critical perspective | Comment critically on the study, reflecting on the things that went well and those that did not, so others can learn from this experience | Abstract: 3  Discussion: 24-27 |

GRIPP2 Checklist from: Staniszewska, S., Brett, J., Simera, I. *et al.* GRIPP2 reporting checklists: tools to improve reporting of patient and public involvement in research. *Res Involv Engagem* **3,** 13 (2017). https://doi.org/10.1186/s40900-017-0062-2
