## Supplementary material for "Identifying gaps in health literacy research through parental participation": REPRISE framework

The REporting guideline for PRIority SEtting of health research (REPRISE) domains and sub-items as related to our study

Key reference, and use of standardised format from Tong, A., Synnot, A., Crowe, S. et al. Reporting guideline for priority setting of health research (REPRISE).  BMC Med Res Methodol 19, 243 (2019). https://doi.org/10.1186/s12874-019-0889-3

| **No** | **Item** | **Descriptor and/or examples from REPRISE** | **Our study** |
| --- | --- | --- | --- |
| A | Context and scope |  |  |
| 1 | Define geographical scope | Global, regional, national, city, local area, institutional/organizational level, health service | national with a local focus of the workshops on the four locations of HELICAP (Magdeburg, Freiburg, Regensburg, Hannover) |
| 2 | Define health area, field, focus | Disease or condition specific, interventions, healthcare delivery, health system | Allergy prevention, allergic diseases (e.g. also allergic asthma, atopic eczema, food allergies), COVID-19 in children with allergies  Since studies recommend confrontation, rather than avoidance, with allergens, there has been a great deal of uncertainty in the population. Uncertainty is also felt during the outbreak of the Sars-COV-2 pandemic. |
| 3 | Define the intended beneficiaries | This may include the general population or a specific population based on demographic (age, gender), clinical (disease, condition), or other characteristics who may benefit from the research | Parents of children with allergies or families at risk of allergies, parents of children with allergies during the COVID-19 pandemic. |
| 4 | Define the target audience of the priorities | Policy makers, funders, researchers, industry or others who have the potential to implement the priorities identified | Researchers and research funders in the field of allergy prevention, (pediatric) physicians, health professionals |
| 5 | Identify the research area | Public health, health services research, clinical research, basic science | Public health research |
| 6 | Identify the type of research questions | Etiology, diagnosis, prevention, treatment (interventions), prognosis, health services, psychosocial, behavioral and social science, economic evaluation, implementation; this may not be pre-defined | Health literacy research, prevention (interventions)  Not predefined.  Priorities generated by participants focussed on accessing and understanding health information and measuring health literacy as it corresponded to the organization of the workshops, but also wider public health context and healthcare access. |
| 7 | Define the time frame | Interim, short-term, long-term priorities, plans to revise and update | These were intended as interim priorities based on the situation of current very limited data. |
| B | Governance and team |  |  |
| 8 | Describe the selection and structure of the leadership and management team | Those responsible for initiating, developing, and guiding the process for priority setting, and examples of structures include; Steering Committee, Advisory Group, Technical Experts | The process was initiated and guided by a task force of members of the HELICAP research group and its cooperation partner (the German asthma/allergy patient organization DAAB e.V.) |
| 9 | Describe the characteristics of the team | Stakeholder group or role, institutional affiliations, country or region, demographics (e.g. age sex), discipline, experience, expertise | Leads of the HELICAP group are were Prof Dr Christian Apfelbacher (Institute of Social Medicine and Health Systems Research, Otto von Guericke University, Magdeburg, Germany) and Prof Dr Eva Maria Bitzer (Department of Public Health and Health Education, University of Education Freiburg, Freiburg, Germany) |
| 10 | Describe any training or experience relevant to conducting priority setting | Consultants or advisors, members with experience or skills relevant to the conducting priority-setting e.g. qualitative methods, surveys, facilitation | Facilitators were chosen for proven experience in conducting qualitative research/research with parents and were experts in the field of health literacy research. The group currently does not have prior experience in conducting a prioritization process. |
| C | Framework for priority setting |  |  |
| 11 | State the framework used (if any) | James Lind Alliance, COHRED, CHNRI, Dialogue Model, no framework (general research priority setting) | This was general research priority setting (no formal framework) but based on the principles of the James Lind Alliance. |
| D | Stakeholders or participants |  |  |
| 12 | Define the inclusion criteria for stakeholders involved in priority-setting | Patients, caregivers, general community, health professionals, researchers, policy makers, non-governmental organizations, government, industry; specific groups including vulnerable and marginalized populations | Primarily, new parents and parents of children with allergies, families at risk of allergies; secondarily, all other individuals related to the use cases of early childhood allergy prevention and COVID-19 in children with allergies |
| 13 | State the strategy or method for identifying and engaging stakeholders | Partnership with organizations, social media, recruitment through hospitals | Recruitment was carried out by the cooperation partner (the German asthma/allergy patient organization DAAB e.V.), via local day care centers and pediatricians as well as via snowballing. |
| 14 | Indicate the number of participants and/or organizations involved | Number of individuals and organizations, include number by stakeholder group | n=55; detailed in participant characteristics |
| 15 | Describe the characteristics of stakeholders | Stakeholder group, demographic characteristics, areas of interest and expertise, discipline, affiliations | Female focus, highly educated;  detailed in participant characteristics |
| 16 | State if reimbursement for participation was provided | Cash, vouchers, certificates, acknowledgement; what purpose e.g. travel, accommodation, honorarium | Refreshments and snacks were provided during the presence workshops and an expense allowance of 20 euros was offered. |
| E | Identification and collection of research priorities |  |  |
| 17 | Describe methods for collecting initial priorities | Methods e.g. Delphi survey, surveys, nominal group technique, interviews, focus groups, meetings, workshops; prioritization e.g. voting, ranking; mode e.g. face-to-face, online; may be informed by evidence e.g. systematic reviews, reviews of guidelines/other documents, health technology assessment | The preparatory phase of the study served to develop relevant topic areas for the workshops.  Research needs/uncertainties were formulated by the participants based on the following questions designed as a framework for the workshops:   - Welcoming the participants, overview of the workshop objecive and procedure, information about healt literacy - Division of all participants into three thematic units, alternating:  1. “Acessing Information” 2. Measuring Health Literacy” 3. “Understanding information”  - Preview of results, summary   Methods used in the workshop: world-café, focus group dicussion  Method that will be used for further priorization: (online) Delphi survey |
| 18 | Describe methods for collating and categorizing priorities | Taxonomy or other framework used to organize, summarise, and aggregate topics or questions | The collating and categorization of the collected items from the workshops was first done in a dyad system followed by…. ?? Kategorien bilden, Redundanz- und Überlappungsanalyse |
| 19 | Describe methods and reasons for modifying (removing, adding, reframing) priorities | Based on scope, clarity, definition, duplication, other criteria | All priorities raised were carefully discussed in the task force in order to understand the meaning, and if they were duplications of other priorities already raised. |
| 20 | Describe methods for refining or translating priorities into research topics or questions | Reviewed by Steering Committee or project team | Translation of the items collected in the workshop into research questions occurred within the task force, he said. First, the individual facilitators reformulated the items of their own units; these were cross-checked and, if necessary, reformulated by other members of the task force in further rounds. |
| 21 | Describe methods for checking whether research questions or topics have been answered | Systematic reviews, evidence mapping, consultation with experts | Drawing on the group's expertise in areas of health literacy research, literature research in various scientific databases |
| 22 | Describe number of research questions or topics | Number of priorities at each stage of the process | Workshops: unlimited  Delphi: approx. 50 |
| F | Prioritization of research topics/questions |  |  |
| 23 | Describe methods and criteria for prioritizing research topics or questions | Methods e.g. Delphi survey, surveys, nominal group technique, interviews, focus groups, meetings, workshops; Prioritization e.g. voting, ranking; Mode e.g. face-to-face, online; Criteria e.g. need, feasibility, novelty, equity | Delphi-method (ongoing, not in manuscript) |
| 24 | State the method or threshold for excluding research topics/questions | Thresholds for ranking scores, proportions, votes; other criteria | Research questions were excluded if they did not address the topic of health literacy and/or were too far from the topics of our research group. |
| G | Output |  |  |
| 25 | State the approach to formulating the research priorities | Area, topic, questions, PICO (population, intervention, comparator, outcome) | Parents'/participants’ needs and uncertainties are diverse.Therefore topics explored were broad and priorities listed as broad areas for further exploration in future workshops. |
| H | Evaluation and feedback |  |  |
| 26 | Describe how the process of prioritization was evaluated | Survey, workshop | The exercise was conducted as a workshop with preparatory/planning activities (preparatory phase). |
| 27 | Describe how priorities were fed back to stakeholders and/or to the public; and how feedback (if received) was addressed and integrated | Public meetings or workshop, newsletters, website, email, online presentations | At the end of the workshop, all results were presented to the participants and open questions were clarified.  Interim results and findings from the preparatory phase can be viewed via the HELICAP homepage and Spotify (podcasts), and the research group is in email contact with the participants/parents. |
| I | Implementation |  |  |
| 28 | Outline the strategy or action plans for implementing priorities | Communication with target audience, via policies and funding | The strategy is to integrate community participation in future local studies as per the feasibility data and stated willingness of local groups to participate, and to collaborate on future research taking into account these stated priorities. |
| 29 | Describe plans, strategies, or suggestions to evaluate impact | Integration in decision-making, funding allocation, review of relevant documents | The impact will become clear if funded projects focussing on the priority areas is secured. The intention is to move forward to Identify the TOP 10 research priorities finally by means of a consensus-workshop. |
| J | Funding and conflict of interest |  |  |
| 30 | State sources of funding | Name sources of funding for the priority-setting exercise; if relevant include the budget and/or cost | This work was funded by the Deutsche Forschungsgemeinschaft (DFG, German Research Foundation) - project number 409800133 |
| 31 | Declare any conflicts or competing interests | State any conflicts of interest that may be at an individual level and/or at a contextual level (e.g. political issues, controversies) that may affect the process, output or implementation. | none |
